# Deviation from Nash Mixed Equilibrium in Repeated Rock-Scissors-Paper Reflect Individual Personality Traits

**DOI:** 10.1101/2023.04.20.23288903

**Authors:** Kensuke Arai, Suma Jacob, Alik S. Widge, Ali Yousefi

## Abstract

Quantification of human behavior in a social context can reveal behavioral variations in a population that can be used to improve screening and targeting accuracy in the development of interventions and biomedical treatments for psychiatric disorders. However, social interaction is difficult to study and quantify in a controlled, reproducible environment. In this research, we describe an experimental framework that utilizes a zero-sum game of repeated Rock-Paper-Scissors played against an artificial intelligence agent to probe social interaction. Human deviation from the Nash Mixed Equilibrium strategy of play, the only guaranteed way to avoid exploitation, can be seen in the sequential dependence of hands. We hypothesize that this deviation represents humans mimicing randomness through constant adjustments of behavior, which we analyze in terms of a set of switching heuristic lag-1 conditional response rules. We quantify and interpret the set of rules subjects are able to utilize as mirroring individual personality traits of the subjects. Subjects in the study also completed the Autism Quotient Abridged survey, and subscores of the social, imagination and routine dimensions were found to be predicted by a combination of behavioral features derived from game play. This work proposes a new approach to quantifying social behavioral traits in individuals affected by psychiatric disorders that manifest in social functioning.

## 1 Introduction

Symptoms of psychiatric illness often manifest simultaneously and on a continuum at many physiological, neurological and cognitive levels. The mechanisms underlying these complex disorders can be multigenic, and similar symptoms may be attributable to different underlying causes. Currently, clinical psychiatric diagnosis is based on a profile of symptoms generated from self-report and assessment by family members, teachers or psychiatrists [1, 2, 3, 4], but biased response in self-report [5], low inter-rater reliability [6, 7, 8] and inherent subjectivity, are known drawbacks to these assessments. Rosenhahn’s famous experiment [9] sensationaly demonstrated the pitfalls of patient self-report and subjective evaluation of behavior being used in diagnosis, drawbacks that persist even under good diagnostic intentions of patient and physician. Researchers are developing new assays that do not rely on subjective report, like auditory responses [10, 11], motion perception [12, 13], cross-modal audio-visual perception [16], spatial temporal perception [17], visual social cue perception [18, 19], observation of repetitive behavior [14] and body positioning [15], to make diagnosis more automatic, objective and data-driven. Because symptoms may overlap despite different underlying conditions, biomarkers in the genome and metabolic products [20, 21], and in resting state brain dynamics [22, 23, 24, 25], are also being sought by researchers. Accurate diagnosis thus depends on a good assessment of symptoms and biomarkers. However, a core symptom of psychiatric disorders, the impairment of social function, is difficult to assess. Full extent of deficient social functioning may only manifest itself in actual complex, reciprocal social interactions [32], which are difficult to reproduce repeatably in a clinical setting as minimally, a human dyad is required. It is unrealistic to pair every test subject against the same partner, and while interaction with a computer can make a task repeatable, reciprocal interactive laboratory tasks are still rare [34]. Neuroecomonic games [33] have been used to investigate decision making tasks in a social setting, but present unique challenges not present in the analysis of isolated or programmatically illicited behaviors. To address these concerns, we propose using an AI-backed interactive competitive neuroeconomic game [35], that allows study of social interaction, and a relevant and succinct analysis and modeling framework that captures a subtle but robust mechanism of adaptive social behavior. Games restrict the degree of freedom of environment and actions that allow easy state description, yet may still exercise some of the same social cognitive skills required in natural social interaction like reading anothers intentions [36, 37]. We argue replacing human opponents with an AI agent is a good surrogate to having the same test opponent paired to every participant, permitting repeatable social experiments with reciprocal interaction across a participant population to be studied. A realistic social experiment would also allow concurrent monitoring of biological signals underlying the quick, that may elucidate how behavior and cognitive process differs in affected individuals by exploring mechanisms inaccessible through behavior alone.

Rock-Paper-Scissors (RPS) is a popular game worldwide often used to settle playground disputes. After one round, the loser might move the goalpost and declare “best of 5”. In longer games of repeated Rock-Paper-Scissors (rRPS), any pattern or bias a player exhibits becomes open to exploitation. The Nash Mixed Equilibrium (NME) in rRPS chooses R, P or S at random with equal probability, and is the only guaranteed strategy that avoids exploitation. Humans are poor generators of random behavior [38, 39], and investigation has focused into how human behavior deviates from the normative standard of the NME. Evidence of deviation initially focused on whether marginal action distribution is biased away from equiprobability, with some investigators finding a slight bias towards R [40, 41, 42], although the results were not consistent across studies [43]. Another potential point of deviation is in the presence of sequential dependence in the moves, and studies have typically looked for one of several commonly considered dependency rules. The most widely studied ruleset is the operant conditioning principle of win-stay-lose-switch, an approximate algorithm for Bayesian reinforcement learning, [44, 45, 46]. Forder *et al* [45] found that the degree of fidelity to win-stay probability is flexible as the reward value is adjusted in non-zero sum versions of rRPS, while the lose-switch probability remains unchanged. Srihaput *et al* [46] found that when playing repeated rounds against a randomly assigned opponent, win-stay probability drops but lose-switch probability remains unchanged when comparing consecutive rounds where the opponent remained the same versus consecutive rounds where opponent was changed. These results suggest different neural pathways that generate the actions following win and lose, which may be an evolutionary behavioristic consequences of the higher risk to the organism of repeating failures than experimenting with conditions leading to success [45, 46]. In an experiment where several 6-player groups played rRPS, players were randomly re-assigned a partner from within their group. A slight, group-level bias for one hand that slowly cycled in order (R*→*P*→*S*→*R*→*P*→*S or reverse) emerged, and the cycling period of each group could be explained by the mean win-stay-lose-switch probabilities of that group, [41], demonstrating an observable consequence of sequential dependency in player moves. Other heuristic rules like cycling and Cournot best response (choose hand that beats last opponent hand) have been considerred, and schizophrenia patients have shown pronounced preference for ascending cycling against computer opponent, and choosing the Cournot best response against human opponent [40]. Most of these studies find the population mean of these probabilities to be near 1/3, which is not a strong indication of serial dependence, yet serially dependent heuristic strategies have been codified by expert players (WRPSA website:”Strategies”) like “win-stay, lose switch”, mimicry (ie copy opponent’s last move or beat opponent’s last move) and cycling. Zhang *et al* [47] has found subpopulation among study participants of those who play win-stay-lose-switch, and those who play win-switch-lose-stay, suggesting not only the importance of considering the individuality of subjects, but also the possibility of personality being reflected in the indiviuality of game play.

A young child might play a string of Rs, and our ability to recognize this and counteract it, is an example of reasoning about the competitor and exploiting a glaring regularity. On the other end of the spectrum, macaques playing against computers programmed to play NME end up playing the same hand over 90% of the time [48], a realization perhaps that there is no regularity in the opponent, and also that the opponent will not exploit even the most obvious regularity. Batzilis *et al* [49] has found that in first-time encounters with an opponent, players are closer to the Nash Mixed Equilibrium, but upon subsequent encounters, use accrued information about them to deviate from the NME, and are able to exploit opponents. Brockbank *et al* [50] has investigated classes of regularities humans can exploit by pairing against computer opponents that were programmed to exhibit sequential regularity, found a wide ranging difference in how many subjects successfuly exploited different rules, but typically advantages were not visible until tens of rounds were played. Stoettinger [51] has found that adapting to a dynamic competitor occurs more efficiently when the competitor changes to structurally similar rules. These results suggest that human players adapt, and are not strictly following any one heuristic, as doing so would open the player up to exploitation. Most studies have limited investigation of sequential dependence to the most recent past round, and have not addressed this adaptation. Brockbank *et al* [43] has found no evidence of win streaks or losses that are expected if adaptation occurred transiently and on a short timescale [52], leading to the hypothesis that humans adapt over long periods to aggregate patterns of play, and considered dependencies that can potentially reach further into the past beyond the last round. We hypothesize that rather than a slow adaptation process, heuristic used in game play changes abruptly on a scale of a few rounds, possibly to mimic randomness. We identified what heuristic was being used at all times during the game, and detected the timing of when heuristics change, and found changes coincide with a sharp regaining of advantage against the AI, whose most recent learned behavior of the human player suddenly becomes obsolete. Individuality in human-generated random sequences [38, 39] and its predictive link to capacity for creative thinking [53] have been reported. In the context of rRPS, we found that the repertoire of rules which a player chooses from is characteristic of each player, and details about the repertoire are predictive of ASD personality traits as measured by the Abridged Autism Quotient answered by each subject. Autism is a potential use case of the rRPS as a social behavioral assay, as deficits in social function is a defining feature of autism, though this behavioral assay may also be sensitive to deficient social function in other disorders.

## 2 METHODS

### 2.1 Experiment

1370 anonymous volunteers (human players, HP) were recruited from the online cognitive experimental platform testmybrain.org [54] between July to December 2021 to play 300-round games of rRPS (cyclic dominance rules rock beats scissors, paper beats rock, scissors beats paper) against an AI agent running on a web browser, playable on iOS, Android, Windows and Macintosh. The AI was comprised of 3-perceptrons each representing R, P and S [55, 56], whose internal parameters were updated online after each round, and that predicted the next HP move using the past 2 HP and AI moves. The rounds were self-paced, and a noticeable degradation of performance was observed for participants who spent less than 600ms per round, leading us to suspect lack of engagement in such participants, and they were subsequently removed from analysis. In addition, participants were also removed if there were more than 10 round-round intervals *>* 1 minute, or more than 15 consecutive repeated responses. In addition to playing the 300-round game, participants also completed the 28-item Abridged Autism Quotient [4] (AQ28), whose composite score is a sum of 4 subscores assessing social behavior - Social Skills (8 items), Imagination (8 items), Routine (3 items), Switch (4 items) and a subscore assessing fascination with Numbers and Patterns (5 items), Fig. 1D. This left 298 players who completed a 300-round game and were deemed credibly engaged most of the time, and a 193-player subset who also responded to the AQ28 survey. We refer to these populations as HPvAI(0) and HPvAI(AQ), respectively. Most of the participants were in their early 20s, with slightly more identifying as males than females, Fig. 1A,B. The mean net wins over a round for these HPs was -15.4, with standard deviation 28.1, 1C. The participant mean AQ28 of our particpant population was 76, which compares to a mean of about 55 for healthy populations and 85 for autistic populations reported by Hoekstra *et al* [4].

**Fig. 1.**
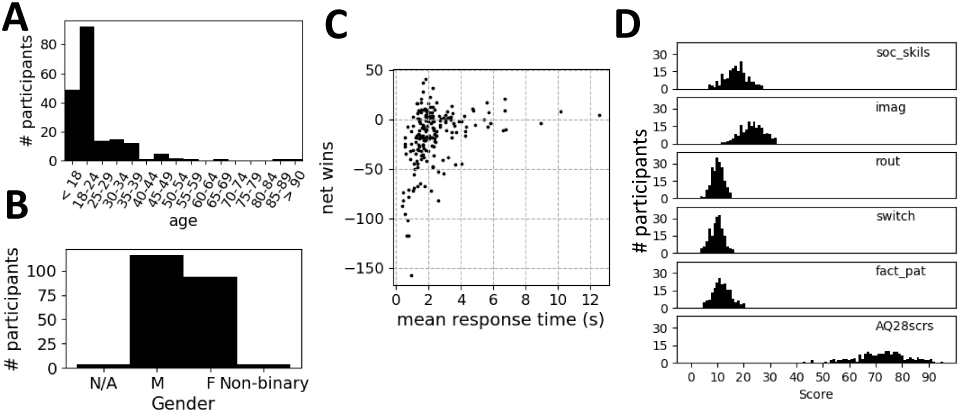
Demographic and basic game statistics of HPvAI(AQ) participants. A) Age and B) gender distribution of participants. C) Net wins versus mean response time (round duration). D) Distribution of Abridged Autism Quotient subscores and composite score. Bottom figure is the sum of top 5 subscores.

### 2.2 Lag-1 rules

Lag-1 conditional response rules are rules of the form “take action Y if condition X occurred in previous round”. In the context of rRPS, we consider 3 natural sets of triggering conditions that occur within the game: 1) HP’s own hand (**R**ock, **P**aper, **S**cissors), or 2) AI’s hands ***r***_A_ock, ***p***_A_aper, ***s***_A_cissors, or 3) the outcome of HP and AI’s hands ***w***in, ***t***ie, ***l***ose. We do not consider finer-grained conditions like “won (tied) (lost) using rock (scissor) (paper)”. HP’s actions that follow can be 1) the next hand itself **R**ock, **P**aper, **S**cissors, or transition with respect to the 2) player’s or 3) AI’s last hand **D**owngrade or **D**_A_owngrade, ie choose move that loses to own or AI’s last hand, **C**opy or **C**_A_opy, ie repeat own or AI’s last hand, or **U**pgrade or **U**_A_pgrade, choose move that beats own or AI’s last hand. By combining a conditions and actions set, a ruleset can be created that generates the next action. For example, combining outcome and transitions from HP’s last hand can result in a deterministic ruleset “**U**pgrade after ***w***in”, “**C**opy after ***t***ie” and “**D**owngrade after ***l***ose”. Or combining AI’s last hand as conditions and directly specify a hand as an action result in a set of rules like “**P**aper after ***r***_A_”, “**P**aper after ***p***_A_” and “**R**ock after ***s***_A_”. This last rule picks a move that beats the AI’s last move, except when AI plays “***p***_A_”, in which case it copies the AI. Because there are 3 actions that can be taken in rRPS, the above rule statements are actually 3 probabilistic rules *p*(R*|r*) = 0*, p*(P*|r*) = 1 and *p*(S*|r*) = 0. For each set of conditions, only 2 of the 3 set of actions describe a unique set of conditional rules. Moreover, DCU*|rps* is just a restatement of RPS*|rps*, and identical moves can be generated using either description. For the remainder of this paper, we choose the following 6 conditional rule sets DCU*|wtl*, RPS*|wtl*, DCU*|rps*, DCU*|r*_A_ *p*_A_ *s*_A_, D_A_C_A_U_A_, D_A_C_A_U_A_ *|r*_A_ *p*_A_ *s*_A_. We call each pairing of condition and action set a framework. Each framework has 3 *×* 3 = 9 set of rules and 6 unique probabilities. We will search for evidence of rule use by humans by testing the 6 *×* 9 = 54 candidates from the lag-1 set of rules.

### 2.3 Detecting Repertoire rules

Response probabilities have been calculated over the entire game without considering possible temporal dynamics, and used to detect changes in behavior across various competitive conditions or behavioral differences between populations in existing research. We explicitly consider the temporal dynamics of this probability as reflecting the switching of rules, and outline its estimation from data. If a particular lag-1 rule (U*|w* to be concrete) is being used at any point of time during the 300-round game, we expect to see an abundance of consecutive wins (not necessarily occurring in consecutive rounds) that are followed by Up transition in the game data. Finding all the wins in the data, define RND(win #*n*) as the round index of the *n*th win. To construct a test to quantify this, we first estimate *p*(U*|w*)(round) using sliding windows of size *B* (=3 win conditions). We estimate *p*(U*|w*)(RND(win #1)) by using the 1st, 2nd and 3rd wins, and calculate 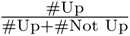 following those wins. Now slide over to the 2nd, 3rd and 4th wins to estimate *p*(U*|w*)(RND(win #2)) using the same procedure. We hypothesize the rule for win persists in the mind even in rounds not matching the “win” condition, so we intrapolate between rounds that were won to obtain *p*(U*|w*) for all rounds. We then count number of rounds where *p*(U*|w*)(round) *>* threshold of 0.8 over the whole game. To check the significance, we repeat this process using data with order of the 300 rounds shuffled to create a null distribution of data with move content intact but with temporal structure destroyed. If the number of rounds above threshold (rule shuffle score, RSS) is 98th percentile or higher of this null distribution, we deem U*|w* to be in the repertoire. Doing this for all 54 lag-1 rules, we determine the set of rules that are in the repertoire.

### 2.4 Detecting Rule Switches

After we find which rules are in-repertoire, we next detect at which rounds switching to a different rule in the repertoire ocurred. As an example of what we expect rule changes to look like, Fig. 4A top trace shows rapid swings in *p*(U*|w*)(round) from nearly 0 to 1 or 1 to 0 around rounds 50, 75, 100, 125, 150, 190, 250, 260. We checked our ability to detect rules changes like this in data, by simulating players that 1) switch between rules shown in Fig. 2B and as a comparison, players that played according to 2) the NME. We refer to the data collected in experiment 1) as RS(n)vAI(i), with n = 1 or 2 specifying which set of rules used, and i = 11, 14 specifying the mean number of rounds between rule switches that are drawn uniformly near the mean) and in experiment 2) as NMEvAI. In RS(n)vAI(i), we know which of the 54 possible rules are in the simulated HPs repertoire, and we know the exact moment rule sets were switched. In the NMEvAI, we know there is no conditional structure in the data, as well as no rule changes.

**Fig. 2.**
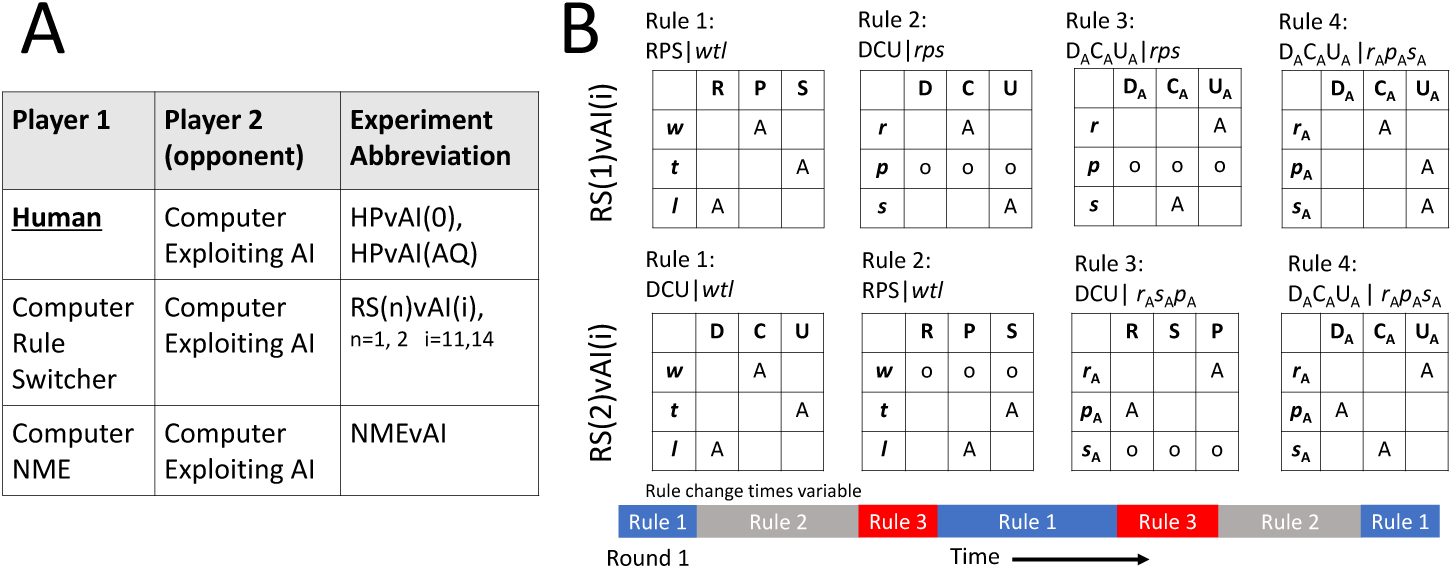
Experiment information. A) List of abbreviations used for experiments and simulations, and the identities of player 1 and 2. Data from human players HPvAI(AQ) ⊂ HPvAI(0). B) Rules used in RS(1)vAI(i) and RS(2)vAI(i) simulations. The conditional probability matrices for each ruleset shown, with A=0.9, empty square=0.05, and o=1/3. Rules where one action nearly deterministic represents repertoire rule, for example *p*(C|*r*) = *A* in Rule 2 of RS(1)vAI(i), while there are a few conditions where all the actions are set to be equiprobable, like the *p*(D,C,U|*p*) = *x* in Rule 2 of RS(1)vAI(i) to simulate a possible case where player showed little serial dependence for that condition. A cartoon of these rules being switched in simulation for RS(n)vAI(i) shown below probability matrices.

### 2.5 Correlation of Behavioral Features to Subscores of Abridged Autism Quotient

The Abridged Autism Quotient is a 28 item scale, with 4 subfactors Social Skills (7 items), Imagination (8 items), Routine (4 items) and Switch (4 items), and Numbers and Patterns (5 items). Each item is scored on a 4 point Likert scale, and the scores from each item for each subfactor are summed, while the composite score is the sum of all 28 items. Bonferonni corrected correlation coefficients between each feature and the subscores were calculated, Fig. 7A. The predictive power these features have of subscores were also investigated using nested cross-validated Lasso. We have little to no preconception as to what features might explain each subscore, and as such we sought an impartial and systematic method to find a sparse set of explanatory features for each subscore. Behavioral features and scores from the SUB population were split by 4-folds. Foreach fold, the cross-validated Lasso using L1-regularization was used to determine optimal regularization parameter *α* and sparse weights for the features using an inner 3-fold train/test split, and this model was used to predict HP subscores using test features. Care was taken to prevent data-leakage between the inner and outer loops. The median *R*^2^ for the prediction of each subfactor was reported across folds for the data, as well as for 100 realizations of shuffled HP IDs. For each HP ID shuffle, *R*^2^ was calculated for all subfactors with the shuffling fixed.

## 3 Results

### 3.1 Reproduction of existing results

Our contribution builds on recent research that addresses the existence of serial dependence of moves [40, 45, 43, 50, 47], an expected feature of non-NME play. We first reproduce results of existing research to show that under the identical metrics, the behavior of our population is in line with previous experiments. Lag-1 conditional responses calculated over the entire game and averaged over the population often do not reveal a strong serial-dependence, as components are very near 1*/*3, as reported by [40, 45], and that we confirm, Fig. 3A. However, differences between responses of healthy and schizophrenic patients [40], and the relative difference in flexibility of components in non-zero sum versions of rRPS [45] or when opponents are changed [46], were detectable using population mean. In agreement with [47], individual responses showed significant heterogeneity in the responses, though agglomerative clustering did not reveal stereotypical patterns of heterogeneity, Fig. 3B, which shows low-dimensional embedding of the 54 components of the conditional responses using t-SNE, with the coloring of the data points corresponding to cluster labels. Compared to a similar analysis of repertoire rules shown in Fig. 4D where clusters are readily visible, clusters are not apparent in conditional responses. Fig. 3C shows the response times (duration of each round) following wins, ties and losses, and reproduces the results obtained by [45] where response after wins is significantly slower than those after ties and losses, suggesting different neural pathways active in decision making. Fig. 3D shows the autocorrelation of wins. [43] reported a similar lack of autocorrelation, arguing against exploitation of opponents using transient move patterns and instead arguing for longer-term adaptation to opponents.

**Fig. 3.**
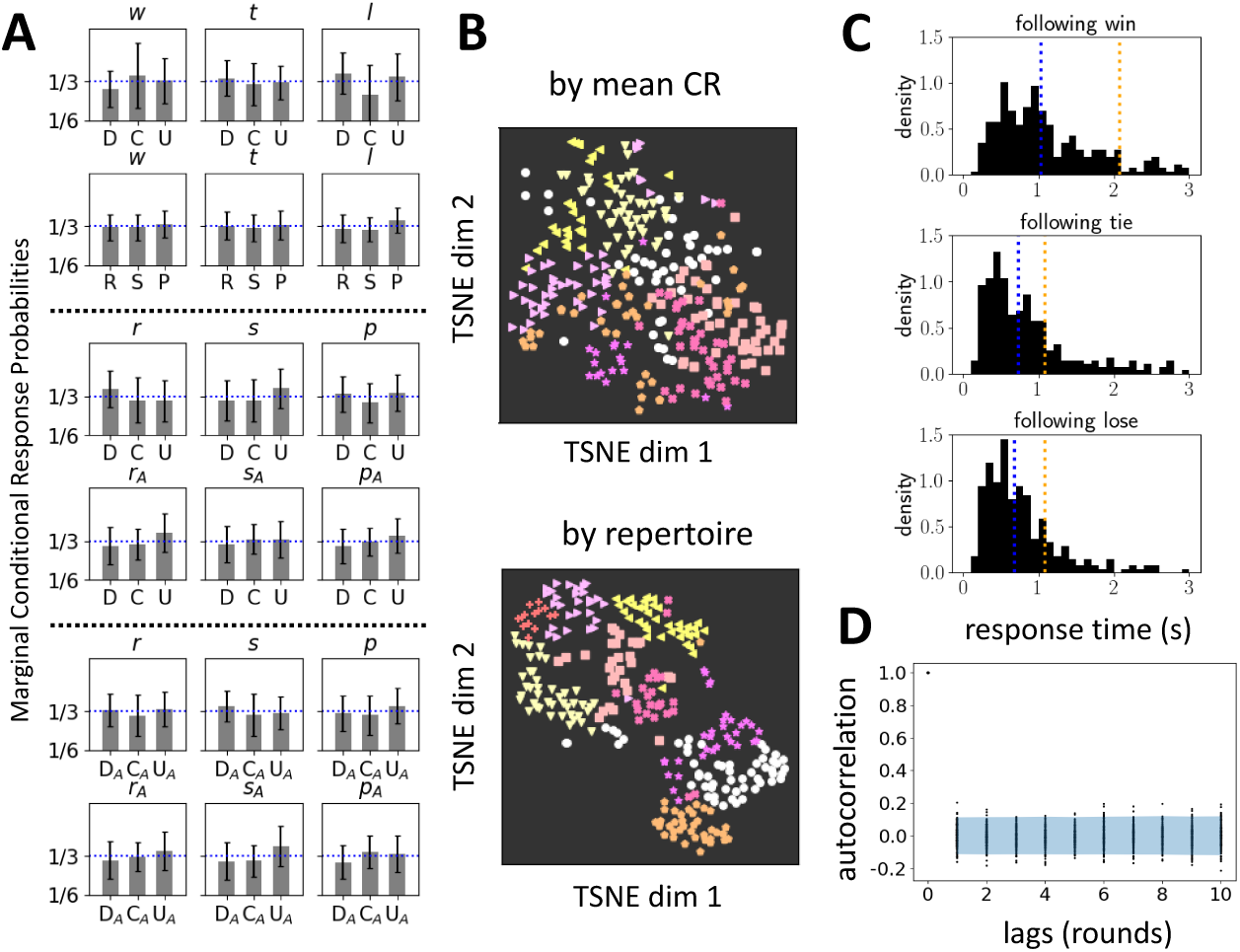
Reproduction of key existing results using HPvAI(0) population. A) Population mean conditional response probabilities calculated over the entire game for each of the 6 frameworks. Conditions indicated in lower-case italic above, actions in upper case below. Most components are near 1*/*3. B) Low-dimensional embedding of the the conditional responses using t-SNE, color-coded by cluster labels obtained by agglomerative clustering, top. The embedded data do not appear clustered in comparison with the embedded RSSs used in detection of in-repertoire rules, reproduced below from Fig. 4D, where clusters in the embedded data are visible. D) Response times after wins, ties and losses. Blue and orange are mean and median, respectively. E) Auto-correlation of the win timeseries. Each point represents lagged auto-correlation for a single HP, and the blue band is the 95% confidence region for null distribution obtained by shuffling round orders.

**Fig. 4.**
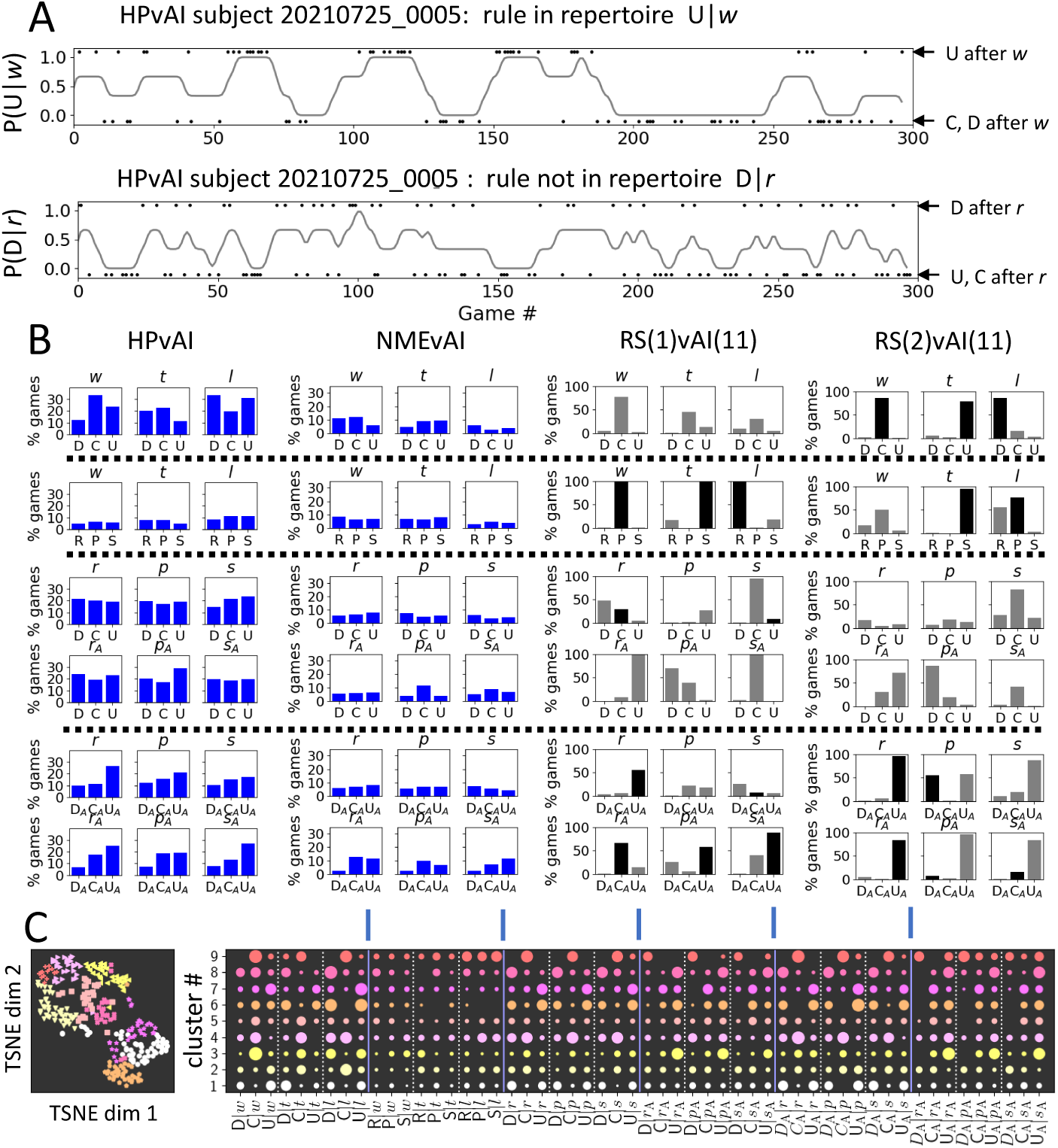
Detection of rule repertoire. A) Example traces of the dynamic conditional response probability calculated for 2 of the possible 54 rules from a HPvAI(28) participant, with top rule classified as in-repertoire. Conditioning events (black dots) plotted at top and bottom represent occurrences that match or do not match the rule, respectively. Rules in-repertoire tend to have interspersed, extended periods where probability is near 1, while rules not in-repertoire rarely go near 1. B) Fraction of the population in which each of the 6 x 9 rules were classified as in-repertoire for experiments HPvAI(0), RS(n)vAI(11) and NMEvAI. Ground truth knowledge of which rules are in the repertoire for RS(n)vAI(11), plotted as a black bar. For the NMEvAI, near chance level detection equally across all rules, as expected. The dotted lines organizes the figures by their transitions to assist in visualizing the pattern present in HPvAI. C) There are stereotypical patterns in the heterogeneity of the repertoire in HPvAI(0). The 54 RSSs used in detection of in-repertoire rules for all the subjects were projected onto a 2-dimensional manifold using t-SNE. Spatial clusters with significant empty space between them in the 2-D distribution of points well matches the colored labels assigned by Agglomerative Clustering set to find 9 clusters, left scatter plot. The pattern of rules that are in-repertoire for each cluster shown on the right. Circle size indicates fraction of subjects in the cluster where rule was in-repertoire.

### 3.2 Rule repertoire detection

Fig. 4B left, middle and right figure compares the fraction of all the data in RS(n)vAI(11), NMEvAI, HPvAI(0), respectively, in which each of the 54 rules was classified as in-repertoire. There is a clear difference in the number of rules in-repertoire detected in the 3 types of data, with the fewest rules in-repertoire detected in NMEvAI, as expected. The identities of the detected in-repertoire rules are also found to be reasonably close to what is actually in the repertoire for RS(n)vAI(11). Also, the probability of any given rule detected as in-repertoire is much smaller and more uniform in NMEvAI, as expected. HPs, unlike the population of simulated rule-switchers that all had identical rule repertoires, presumably have their own individual repertoires, and hence the fraction of any rule being in-repertoire is much lower in HPvAI(0) population than RS(n)vAI(11) simulated population.

There is clearly structure present in Fig. 4B for HPvAI(0) that is potentially informative about the cognitive process of human competition in rRPS. The C*|w* component is often in-repertoire, while the C*|l* component seldom is. This result is reminiscent to that found by [45, 46] that showed the C*|l* component being flexible in different competitive conditions. The 2nd row shows none of the rules from RPS*|wtl* have a high probability being in-repertoire in HPs, suggesting that HPs likely think of moves in terms of transitions. This is reasonable considering the cyclic dominance of the RPS hands. The 3rd and 4th rows of Fig. 5B show transitions relative to HPs own hands. We see the fraction of transitions D,C,U that are in-repertoire, to be about equal to each other across all trigger conditions. This is reasonable since there should be no preference for any transition direction relative to HP’s own last hand under cyclic dominance. In the 5th and 6th rows, D_A_C_A_U_A_ are transitions with respect to AI’s last hands, and correspond to the least, 2nd and best Cournot responses [40, 43]. Interestingly, in every condition, the rank of the frequency of the rule being in-repertoire are the same as its Cournot response rank.

**Fig. 5.**
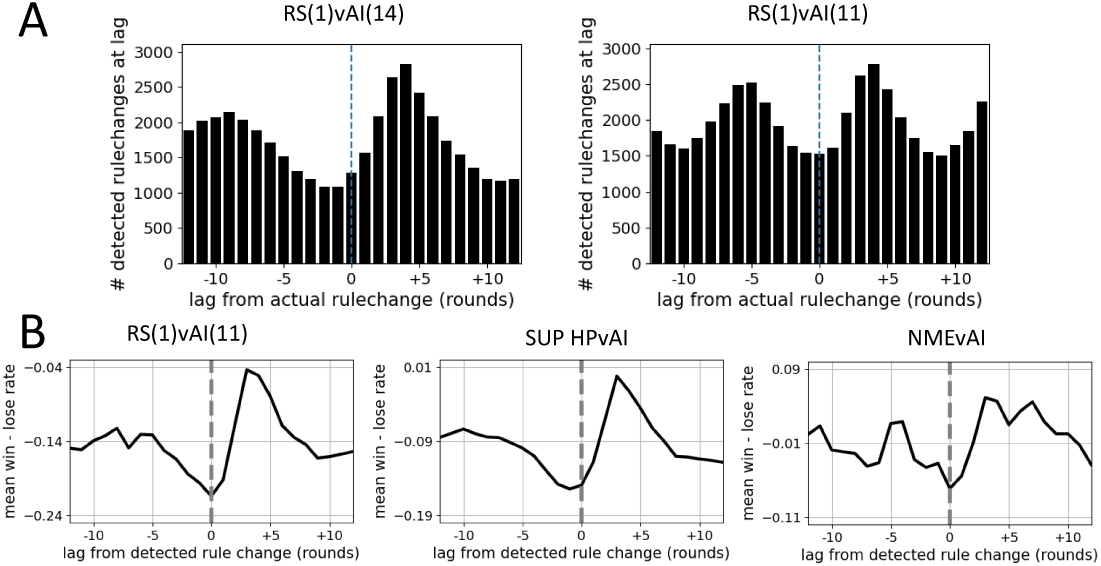
The detection of rule changes and the regaining of advantage. A) Histogram of detected rule change times triggered by ground truth rule change times. The peak is not at lag 0 because rules for each of 3 conditions are changed simultaneousy in simulation, but effect of change is not detectable until the condtioning event occurs in data, resulting in an offset of about 4 rounds. Wide peaks indicate the rule change detection method produces many false positives. B) Consequences of rule-change. RSTW for HPvAI(0), RS(n)vAI(i), and NMEvAI, were calculated by searching for rule changes in 298, 300, and 300 games, respectively. Prominent increase in wins occurs near instances of detected rule changes in HPvAI(0) and RS(1)vAI(11) (RS(2)vAI(11) similar, not shown) but much less in NMEvAI, as expected. Rule changes are detected in NMEvAI, but in a much smaller fraction of games (100%, 94% and 34% of the games, respectively), and even in games where detected, the number of rule change events themselves are significantly smaller as well (80, 38, 3 per game, respectively). Consequently, the number of triggering events used in RSTW for NMEvAI is an order of magnitude smaller than HPvAI(0) or RS(1)vAI(11). The shape of the RSTW is not only a reflection of rule changing, but the AI continually learning the regularities present in HP. Pitting the rule-switcher of RS(1)vAI(11) againt an NME, a similar number of rule changes are detected. However, the RSTW is flat, without the characteristic dip prior and jump after 0 seen for HPvAI(0) and RS(1)vAI(11) (result not shown) because the NME does not adapt to an opponent’s regularities.

Fig. 4B shows that at the population level, a *post hoc* justification for which of the rules are often found to be in-repertoire can be made from considerations of the rules of RPS. At the same time, it is likely that there are considerable individual variations in what is actually in each HP’s repertoire. Fig. 4C shows clustering of pattern of in-repertoire rules by using the percentile of time spent above threshold for each conditional probability timeseries, calculated for each framework as features. This data was projected onto a low-dimensional manifold, where clear spatial clusters were visible. These clusters generally coincided with labels obtained from Agglomerative Clustering of the same features, suggesting that the pattern of in-repertoire rules fits one of a few stereotypical patterns. Fig. 4C bottom row for cluster 8 for DCU*|wtl* show a contrarian group of HPs for whom C*|l* is in-repertoire, and cluster 7 shows a group who do not use any C rules in-repertoire. D_A_C_A_U_A_ transitions show groups who switch the best Cournot rules, and other groups who switch the 2nd best Cournot rules.

### 3.3 Rule change detection

To test our ability to detect rule-switch times from data, we compared the known rule-switch times in the simulated RS(1)vAI(i) data to the detected rule-switch times, Fig. 5. Though we found a large number of false positive detections, Fig. 5, both the rule change times and the interval between rule changes were also detected correctly, both important features of the observed rule-switching. The current work seeks to demonstrate the plausiblity of the rule-switch hypothesis and narrow down the model class for future work that would improve upon the current detection method.

### 3.4 Advantage regained at rule change

An expected consequence of episodic rule changing employed against an opponent constantly looking to exploit, is that advantage is temporarily regained immediately after a rule change. Because the AI is constantly updating its internal model of the HP, sudden changes in HP behavior should result in the internal model of the AI becoming obsolete. We define the rule-switch triggered net wins (RSTW), constructed by stacking the segments of the timeseries of outcomes preceding and following detected rule switches. At each lag from the triggering rule switch, the number of losses is subtracted from the number of wins at that lag, is divided by the combined number of wins and losses, RSTW(lag) = #wins at lag *−* #losses at lag)*/*(#wins at lag + #losses at lag). Fig. 5A right shows RSTW abruptly increasing at moment of rule change, as expected of a cat and mouse dynamic. Brockbank *et al* [57] looked at the autocorrelation of wins, Fig. 3, butdid not see evidence of streaks of wins and losses, which may have been because of human dyads being used. AI plays consistently throughout the game, likely allowing the consequences of rule-switching to be visualized through the RSTW. We emphasize here that the RSTW is not only a reflection of rule-switching, but also the constant learning of the AI. Pitting the same rule-switcher used in RS(1)vAI(11) against an NME will result in rule switches being detected at the same rate as in RS(1)vAI(11), yet the RSTW will be flat.

### 3.5 Deriving features from RPSvAI game data

We have presented evidence that the time-dependent conditional response probabilities is a representation of social cognition underlying competitive rRPS, and we have shown that rule-change points can be inferred from them. We hypothesize that subtle behavioral features that reflect the individuality and personality of the HPs can be numerically derived from our data, and that they can potentially be used to differentiate behavior between healthy and psychiatric patients, can be derived from them. We define candidate features that capture the A) rule repertoire and B) the interplay between the rules in the repertoire. Because the AI is constantly attuned to the most recent moves and the regularities present in them, we looked to C) the internal state of the AI as another source of objective behavioral features. Finally, we included D) conditional outcomes following *wtl* as an easily measurable feature of the possible difference in decision-making pathways of the win and lose condition.

More concretely, A) are the standard deviation of the conditional probabilitiess (6 frameworks x 9 rules = 54 features) and B) are the correlation coefficient between pairs of rule conditional probabilities (54 x 53 / 2 features). We grouped these correlations into terms from 1) same framework, same condition (ie p(D*|w*) and p(U*|w*)), 2) same framework, different condition, (ie p(U*|w*) and p(U*|t*)), 3) different framework sharing condition class, same condition, (ie p(U*|w*) and p(R*|w*)) 4) different framework sharing condition class, different condition, (ie p(U*|w*) and p(R*|t*)) and 5) different framework that do not share condition class (ie p(U*|w*) and p(R*|s*_A_). We did not account for whether either rule was in-repertoire or not. However for this analysis, we deemed it sufficient. For C), the AI’s internal state is not easy to interpret, but the relative magnitudes of the 3 outputs of the perceptron can be interpreted as confidence in the decisions. We looked at the decision confidence of the AI perceptrons following *wtl*, *rps*, *r*_A_ *p*_A_ *s*_A_ (3 x 3 x 3 = 27 features), and the conditional outcome (3 x 3 = 9 features) for a total of 1521 features.

### 3.6 Stability of the measured features

If the candidate features we extracted do reflect the individuality and personlity of HPs, we expect them to be stable under repeated measurements. In the HPvAI(AQ) dataset, 15 participants played more than one 300-round game of RPSvAI, and the games were spaced apart on average by about 10 minutes. While this is insufficient for a complete investigation of feature stability, we use these data to find preliminary evidence that behavioral features are not noise and that they have a degree of stability in repeated measurement. For each feature, we calculated the correlation between the 15 values of the measurement from the first 300-round game and the measurement from the second 300-round game, Fig. 6. The features were categorized by how they were derived, and we computed a mean correlation for all the features within that category, Fig. 6A. For each feature, we also calculated correlations between the first 300-round game with the features from the second 300-round game of a randomly chosen subject from with in the 15 subjects. The shuffling was repeated 200 times to obtain a null-distribution of correlations.

**Fig. 6.**
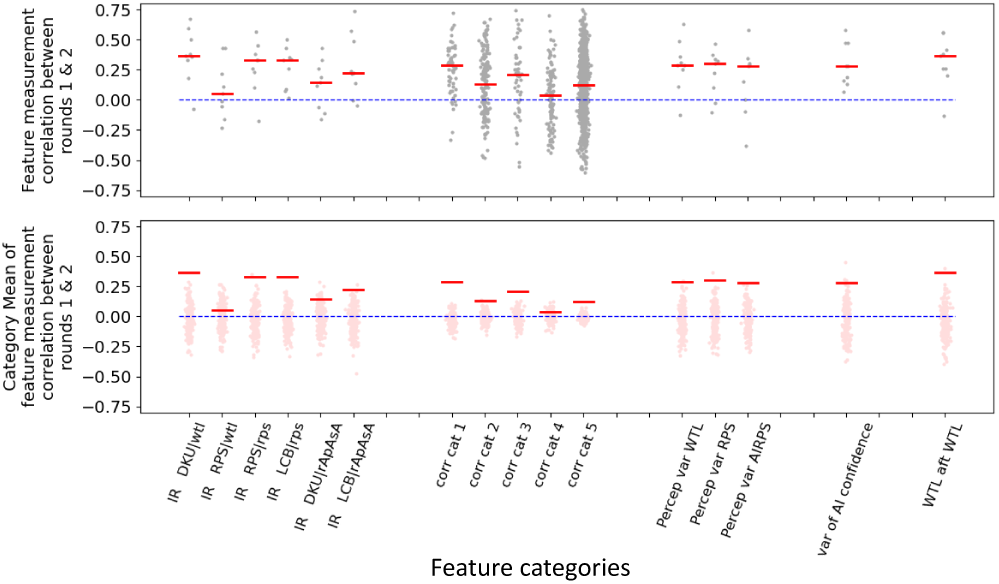
Correlation between the 1st and 2nd measurement of features in HPSvAI SUB. Features systematically generated and organized into categories measured from 15 subjects who played two 300-round games. Top plot shows the correlation between the 1st and 2nd measurement for features in each category. Red line is the mean correlation for all features in the category. Bottom plot shows the mean correlation for different shuffles of the subject index of the 2nd measurement(pink), while red is the same unshuffled mean copied from top plot. Blue line in both top and bottom is 0.

While the correlations are not large, Fig. 6, we are unaware of other reports of stability of a behavioral feature derived from game data. Also, our features were systematically constructed, and it is likely that not all such features are stable behavioral features. It may also be possible that 300 rounds is not long enough to accurately capture some features, especially how certain rules are coordinated with others. Most players were able to complete a round in under 7 minutes, and longer rounds or possibly multiple rounds are a possibility. Also, better methodology used to find in-repertoire rules and detect rule changes, is an obvious area where improvements are possible that may lead to better quality in the extracted features.

### 3.7 Game play features reflect player personality traits

RPSvAI game data features are significantly correlated to and have predictive power for individual AQ28 and its subfactors, the self-reported measure of personality traits of autistic individuals, Fig. 7A. The magnitude of all 1521 correlations were sorted and compared to the null hypothesis to show that there is a relative abundance of large correlations between the features and Social Skills, Routine and Composite, even if they are not necessarily significant after Bonferronni correction. Fig. 7C shows results of the test for predictive power these features have for the subfactors. The mean coefficient of determination *R*^2^ over all folds of the cross-validated Lasso models for each subfactor shown, with the mean *R*^2^ values shown in blue. The means were *>* 0 for Social Skills, Imagination, Routine and the Composite Score, and significantly different than the the null distribution of mean *R*^2^ for the data where participant ID was shuffled (100 shuffles), shown in grey.

**Fig. 7.**
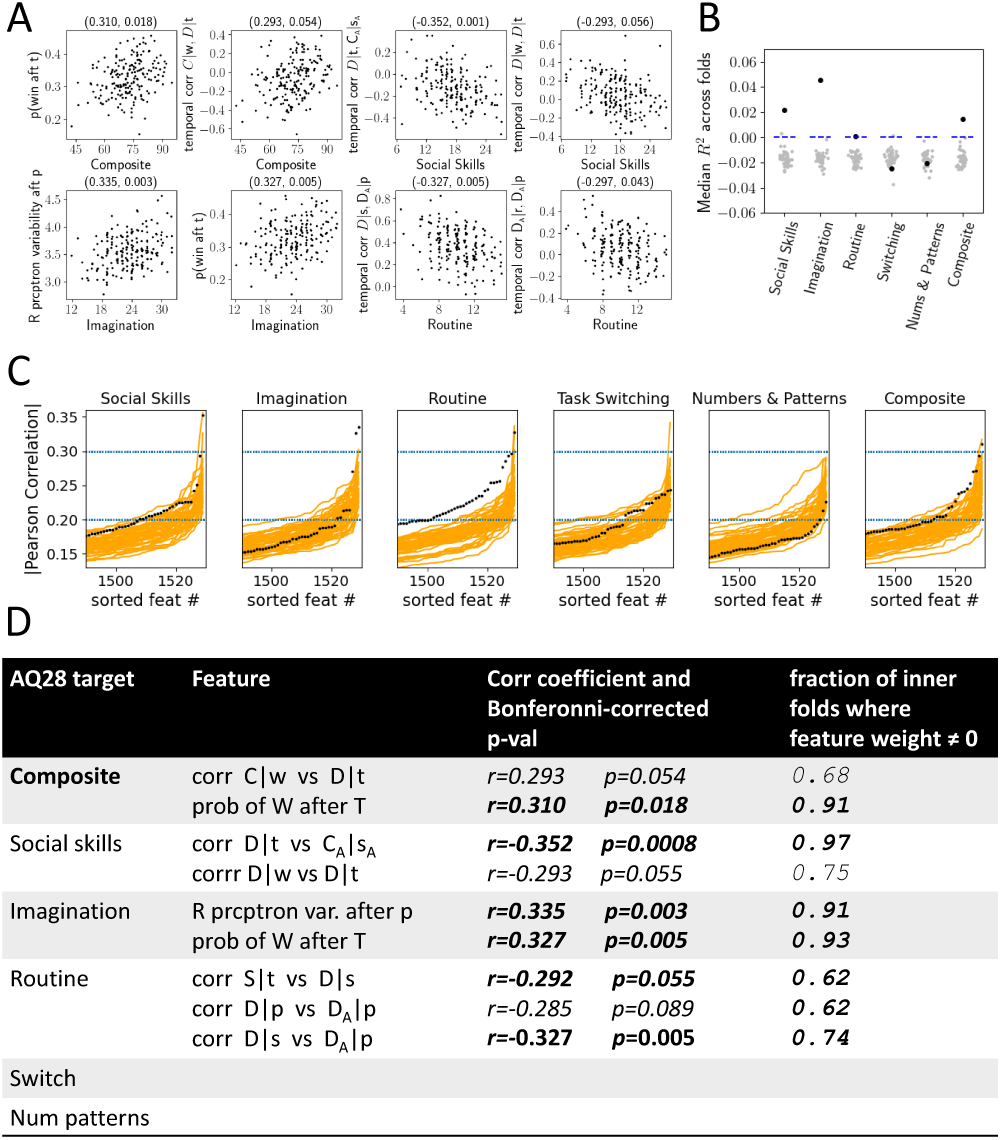
Relationship between features and subscores in HPvAI(AQ) population. A) Scatter plot of features and subfactors whose correlations *r* are significant following Bonferronni-correction. B) The sorted magnitudes of all 1521 *r*s (only top 121 shown) between each of the 5 subfactors and 1 composite scores and the features (black). Orange lines show top 121 *r*s between the subfactors and composite score and the features, but where the HP identities of the features are shuffled 50 times. For Social Skills, Routine, Task Switching and Composite, there are a reltive abundance of features with large correlations with the respective subscores. C) Mean *R*^2^ scores across folds of predictive models of each subfactor (black), and the mean scores across folds for predictive models with shuffled participant IDs (grey). D) List of features with large correlations with subfactors, the corresponding Bonferronni-corrected *p*-values, and the fraction of folds where Lasso weights were non-zero.

The *R*^2^ can take a maximum value of 1, and also arbitrarily large negative values as well, while *R*^2^ = 0 means the mean value of the subscores is correctly predicted. The predictability of the shuffled data are almost always *<* 0, that is shuffled data are not able to predict the mean value of the subscores. Table of features with large magnitude of correlation with subfactors, and the corresponding fraction of folds in which those features had nonzero weight in cross-validated Lasso model is shown in Fig. 7D. The analysis suggests rRPS features are most related to an affinity a subject has to social situations, the ability to read the intentions of others, and whether a subject prefers spontaneity. The rRPS features do not seem to be correlated with a subject’s preference prefer multitasking over concentration on a single task, or a subject’s attention to numbers and patterns seen in the environment. We also note that in none of the shuffles did we observe a high *R*^2^ in multiple subfactors at once, as seen in the unshuffled data. We had very little intuition about which features might correlate with AQ28 and the subscores, and even with the analysis results, interpreting the results is challenging. However, this is not necessarily a negative aspect of this experiment, as it would be very difficult for a patient to guess what the diagnostic goal of playing rRPS is, let alone understanding and invoking behavior that would support a desired diagnosis.

Our *R*^2^ prediction scores for AQ28 and its subscores appear small in comparison to studies showing 70-80% diagnostic accuracy of ASD [24, 15] using brain signal or video analysis. There are several reasons for this apparent poor performance. First, the goal of this study is not to present a computer-assisted diagnosis solution, but to demonstrate a largely unexplored behavioral domain as a source for behavior that potentially differentiates healthy and psychiatric patients. Second, our population was recruited anonymously online, and is not a clinical population with matched controls, resulting in poor sampling of the full range AQ28 and its subscores that such a population would likely provide. Third, subjects performed the AIvRPS experiment without any supervision. A fairly uniform time between rounds suggests some participants concentrated during the task, but a significant number of partipants exhibited a few long pauses between rounds, suggesting they may have had distractions during the task. Such players also tended to lose more against the AI. Because the HP generally displays dependencies on the previous round, such lengthy pauses are expected to result in poorer quality of behavioral features being extracted as the player is likely to not remember what happened in the previous round after such long pauses. Performing the task in a clinical setting should allow better quality data collection.

## 4 CONCLUSION

Humans approximate randomness by using the universal mechanism of rule switching in games of rRPS, which is a specific model of how we deviate from the Nash Mixed Equilibrium. There is heterogeneity among individuals about which sets of rules an individual has available and how they are used together. The choice of rules does not follow from rational consideration of the rules of the game, but better reflect what we know as personality traits. These personality traits translate into differences in measurable behavioral features of rRPS, and we have demonstrated its potential applicablility to the development of standardized diagnostic test for psychiatric illnesses, and to provide a social behavioral biomarker that can be integral part of a joint biomarker together with genetic, neural and metabolic biomarkers. Because this task can be performed in a clinical setting, it is also compatible with concurrent monitoring of biosignals such as heartbeat and EEG, and may allow development of neural biomarkers in the context of social cognitive processing beyond those currently being developed in the resting state or in responses to static sensory stimuli. We believe this will facillitate the link between cognitive behavior that differrentiates psychiatric illnes to brain and physiological activity, possibly opening up avenues for mechanistic investigation of disease.

For future work, we hope to build on our discovery of the rule switching mechanism with improvements in analysis technique. We currently detect rules in repertoire and detect switch times with a plausibe but *ad hoc* procedure. We can use this insight to inform a better selection of model class for a latent-state switching statistical model [58] will be far less prone to noise and false discovery, that will allow us to capture the switching behavior with improved accurracy. We also plan to replace the current rRPS AI with one that better suits the needs of this task. The exploiting AI is crucial in eliciting differentiating behavior, and the 3-perceptron AI has served this purpose, but it is difficult to interpret what the AI is “thinking”. We have found an AI in a kaggle competition (https://www.kaggle.com/competitions/rock-paper-scissors/discussion/221512) that has interpretable states that are similar to our notion of repertoire rules, and may allow us to derive more predictive features from the AI internal state. Replacing our current AI may allow the interpretation of rules chosen by HP in terms of the AI’s internal state, and give clues as to whether rule switches serve to subjective randomness or adversarial reasoning [43]. Being able to elicit behavior that can differentiate healthy and affected states in the laboratory opens up the possibility of observing differences in brain activity. Our work further pinpoints change points in cognitive behavior invisible to the naked eye. Concurrent EEG measurement and employing analysis techniques like global coherence to quantify functional connectivity, is an addition we are currently exploring that may allow us to link neural activity and moment-by-moment changes in cognition [59] that may reveal cognitive differences of healthy and affected individuals. Finally, we are beginning data collection from a clinical population to observe whether the predictability of AQ scores extends to diagnosed population, and rRPS can differentiate healthy and affected individuals.

## Data Availability

All data produced in the present study are available upon reasonable request to the authors

## Supplementary information

Code and data available on github.com, https://github.com/AraiKensuke/AIiRPS.

## Acknowledgements

We than Professors Uri Eden and Mark Kramer for helpful discussions, Shigeru Shinomoto for useful discussion and for providing rock-paper-scissors web code. Sam student. Early volunteer testers of webcode Neurable, RPI, Mayumi Ban, Ayako Nishida, Nanoha Nishida, Hana Nishida.

## Declarations

Some journals require declarations to be submitted in a standardised format. Please check the Instructions for Authors of the journal to which you are submitting to see if you need to complete this section. If yes, your manuscript must contain the following sections under the heading ‘Declarations’:

- Funding
- Conflict of interest/Competing interests (check journal-specific guidelines for which heading to use)
- Ethics approval and consent to participate
- Consent for publication
- Data availability
- Materials availability
- Code availability
- Author contribution

If any of the sections are not relevant to your manuscript, please include the heading and write ‘Not applicable’ for that section.

Editorial Policies for:

Springer journals and proceedings: https://www.springer.com/gp/editorial-policies

Nature Portfolio journals: https://www.nature.com/nature-research/editorial-policies

*Scientific Reports*: https://www.nature.com/srep/journal-policies/editorial-policies

BMC journals: https://www.biomedcentral.com/getpublished/editorial-policies

## Notes

### Competing Interest Statement

The authors have declared no competing interest.

### Funding Statement

Worcester Polytechnic Institute TRIAD grant

### Author Declarations

Institutional Review Board of Worcester Polytechnic Institute gave ethical approval for this work.

### Summary of Updates

1) Added new data previously unused (participants who played game yet did not answer Autism Quotient 28 questionnaire) to add more data to calculation of advantage gain at rule changes 2) Cleaned up and re-organized notation. This allowed us to see clear pattern in rule-use by framework 3) Lower-dimensional embedding using TSNE + clustering analysis shows several stereotypical rule-repertoires in our player population

